# Retention and the Intersection of Structural Inequities in a Breastfeeding Intervention Study

**DOI:** 10.1101/2024.03.15.24304355

**Authors:** Helen Wilde LaPlant, Confidence Francis-Edoziuno, Zhe Guan, Tumilara Aderibigbe, Xiaolin Chang, Ashwag Saad Alhabodal, Kristen Delaney, Dana Scott, Mary Marshall-Crim, Idelisa Freytes, Wendy A. Henderson, Stephen Walsh, Ruth F. Lucas

## Abstract

**Introduction:** Women below the poverty threshold have lower representation and retention in breastfeeding studies.

**Methods:** A secondary analysis of a longitudinal randomized controlled self-management for breast and nipple pain during breastfeeding study. Participants completed online surveys at discharge, weeks 1, 2, 3, 6, 9, 12, 18, and 24, with face-to-face interviews at 6 and 24 weeks. Text messages were sent to participants when modules and surveys were due. Retention was assessed in R with descriptive statistics, Mann-Whitney, Pearson’s chi-square, and Cox Proportional Hazard Regression.

**Results:** Two hundred and forty-four women (89 ≤$50,000 and 155 >$50,000) were recruited. Retention rates at 1 (93%), 2 (87%), 6 (82%), 9 (77%) and 24 (72%) weeks. For women of low income compared to those of high income there was a hazard ratio (HR) of 2.5 (p=0.0001) for retention. For non-Hispanic Black and Hispanic women compared to the combined non-Hispanic White and Other group, HRs for retention were 3.3 and 2.6 respectively (p=0.0001). Adjustment for age in the final hazard regression model of income, age, race and ethnicity decreased the HR for women of low income to 1.6 and HRs for non-Hispanic Black and Hispanic women to 2.1 and 1.9, respectively (p=.0001). However, none of the individual factors in the model achieved statistical significance.

**Discussion:** Retention in breastfeeding studies impacts breastfeeding duration, a key lifelong preventative health behavior. Despite accessible study design, retention of women desiring to breastfeed was adversely affected by the intersection of income, race and ethnicity, and age.

---

Breastfeeding (BF) is fundamental to lifelong protective health equity, yet women and infants below the poverty threshold disproportionately do not exclusively breastfeed for six months (24 weeks), and thus do not receive this benefit.^1–4^ In 2020, 83.1% of women in the United States (US) initiated BF, and 58.2% continued until 6 months.^4^ However, for women below the federal poverty threshold, only 75.4 % ever breastfed and 44.2% breastfed to 6 months.^4^

Despite improvement in overall BF outcomes,^5^ significant disparities in BF rates exist due to social and structural inequities of income, race, and environment. ^2,6,7^ Low income has been shown to have a considerable impact on ability to engage in general protective health behaviors.^8,9^ As 19.5% of Black women and 17.1% of Hispanic women are below the poverty threshold, they are less likely to have health insurance and access to BF resources,^10–13^ putting them at risk of lower BF rates. Cultural, social, and economic factors, such as inequitable access to lactation resources and support, coupled with inadequate knowledge of the benefits of BF, negatively impact BF practice.^14–21^ Therefore, strategies to increase access to BF services and reduce the effects of poverty and structural inequities are necessary for BF equity.^6,18,22,23^

A potential strategy for reducing BF disparities is to increase research study participation of underrepresented groups and individuals at risk of suboptimal health outcomes.^24^ Research outcomes provide evidence that supports the development of disparity-reducing policies and health interventions.^25^ For research and interventions to be beneficial and generalizable, a diverse participant population that reflects the intended user population should be recruited.^26,27^ Effective recruitment and retention strategies contribute significantly to the overall success of clinical studies.^28^ However, recruitment and retention of persons in low-income groups is challenging, resulting in under representation in clinical research studies.^26,28,29^ A text-based intervention is an accessible means of promoting BF equity.^30^ Through the government Lifeline Assistance program, low-income mothers can access smartphones.^31,32^ which enables utilization of text-based interventions to promote BF equity. Therefore, the purpose of this paper is to describe retention rates for women above and below the poverty threshold in a BF study.

## Methods

The *Promoting Self-Management of Breast and Nipple Pain with Biomarkers and Technology for Breastfeeding Women* (*PROMPT*) (R56NR020041, PI, R. Lucas) study took place in partnership with two academic hospitals in the Northeast. The randomized clinical trial tested the efficacy of a text-message and cloud-based patient-informed self-management intervention. The intervention delivers BF information and support through nurse led text-based communication to decrease breast and nipple pain and increase BF self-efficacy and adaptive coping behaviors (Figure 1). For equitable access, text links were delivered via smartphone, which 95% of women of childbearing age possess.^33^ The design used a one-finger swipe method, so women may access the intervention during BF. Women were randomized to intervention or attention control groups and followed via surveys and face-to-face interviews over a 24-week period.

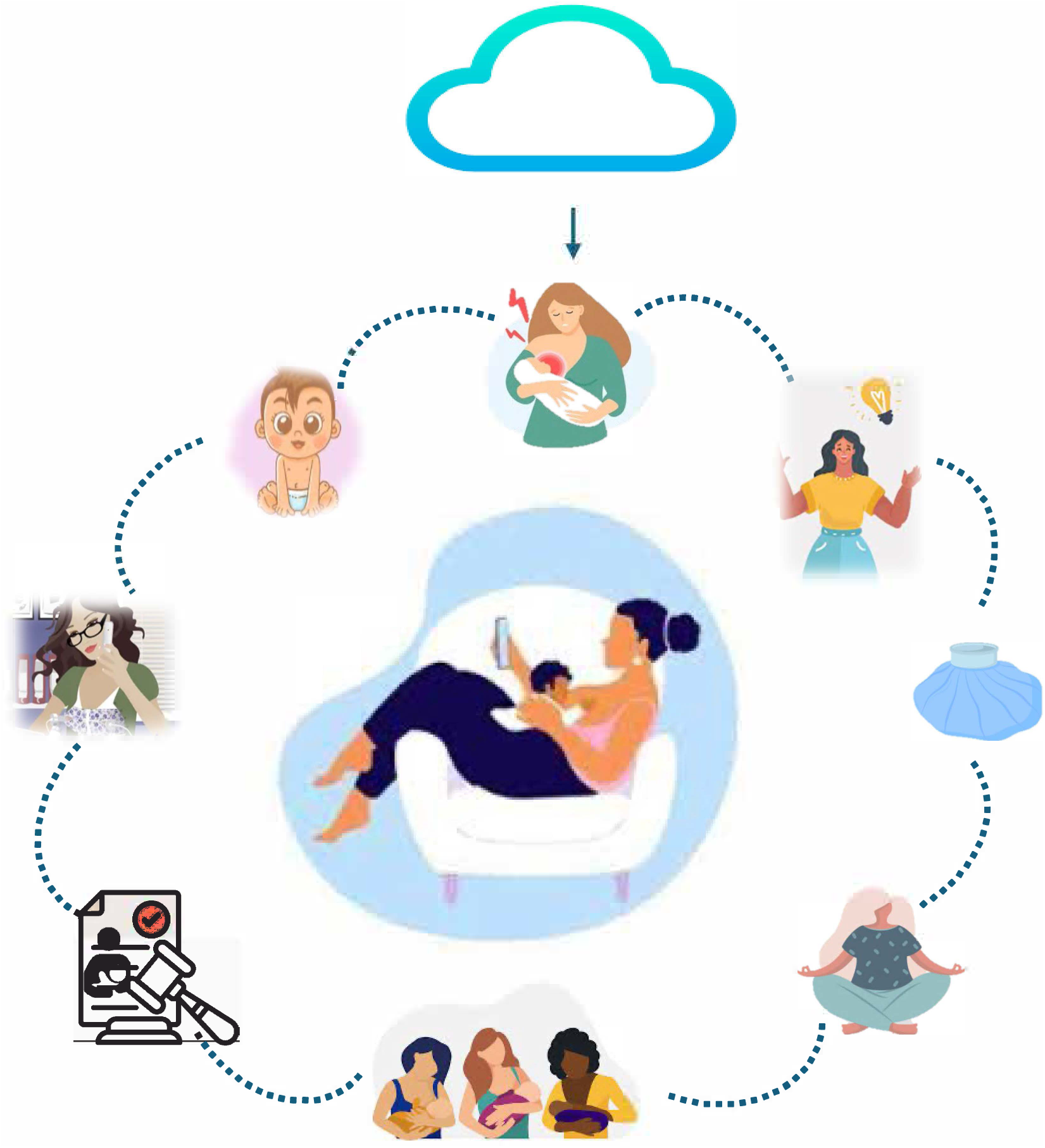

### Sampling

The study aimed to enroll 222 participants. The inclusion criteria were (a) 18 - 45 years of age; (b) birth < 48 hours to a singleton infant of > 37 weeks gestational age; (c) intention to breastfeed; (d) received BF basics during antenatal care; (e) access to the internet via own or study provided smartphone; (f) ability to read and write English; (g) lactation consultant or counselor BF assessment. Exclusion criteria were (a) < 18 years of age; (b) history of significant mental health disorder (e.g., major depression or bipolar disorder); (c) skin conditions on non-dominant forearm (related to pain sensitivity testing),^34,35^ and (d) delivery of an infant with medical complications or congenital anomalies.

### Participant Recruitment

To achieve a diverse sample of participants, members of the research team were hired to represent the ethnic and racial diversity of the women who gave birth at each hospital. The recruitment flyer and educational modules were reviewed by the team for language level and inclusive images. Advertisement was via the PI’s online lab webpage and flyers provided at prenatal clinics and labor and delivery. Screening for eligible participants occurred 4-6 times per week through review of the postpartum census by the project manager in consultation with the PI. The research team prioritized whom to approach first, based on time from delivery, race and ethnicity, and confirmed eligibility with the nurse providing care for each potential participant. Eligible women were approached; if the woman expressed interest, a study flyer was left for her review and a time for return was agreed upon. The research member commonly returned multiple times due to ongoing clinical and infant care, occasionally the next day before the woman was discharged.

### Participant Consent

Eligible women were consented in their private hospital rooms. During enrollment, each participant was assessed for capacity to provide consent, HIPAA consent was obtained, the consent form was reviewed, and the woman was asked three questions to confirm understanding. Screening was documented via REDCap (Research Electronic Data Capture, v.13), a secure web-based database, and consent documents were stored in the clinical setting. Women were randomized using an established dynamic minimization algorithm based on six stratifying characteristics (age, race, route of delivery, BF experience, expected BF duration, and intent to return to work) collected during screening.^36,37^

### Participant Retention

After consent, research team members initiated encrypted text-linked surveys via Twilio (a text distribution platform embedded in REDCap) and confirmed participant receipt and completion. In addition, women completed surveys on BF, pain, and maternal wellbeing and underwent quantitative sensory testing. At enrollment, women without a smartphone were provided a study phone with unlimited data to access the 24-week study.

Following discharge, participants completed online surveys at 1, 2, 3, 6, 9, 12, 18, and 24 weeks and received modules on managing BF pain and challenges, postpartum care, infant development, and maternal mental health. Participants returned at 6 and 24 weeks for in-person interviews and quantitative sensory testing at one of three locations easily accessible by public transport. Scheduling was flexible to accommodate participant availability.

Surveys and modules were sent to participants at predetermined intervals via text links. If items were not completed within six hours, a second invitation link was automatically generated. The online platform allowed participants to begin survey completion and save their results to resume later. Completion of survey instruments and modules was monitored daily by the project manager. If items were not being completed within the one-week window, links were re-sent to participants for remaining surveys or modules in the series, with reminder text messages, telephone calls, and lastly, email messages if participants did not respond to previous methods.

Retention was encouraged through progressive monetary incentives, with a bonus upon completion of the study. To maintain engagement, participants received text links to normal infant development and age-appropriate play modules at weeks 4, 8, 12, 16, 20, and 24. At week 7, a text message was sent to encourage continued participation regardless of feeding method.

### Data Analysis and Management

All data were entered directly into REDCap, a password-protected database, and downloaded for analysis using R software.^38^ Descriptive statistics (percentages for categorical data and means/standard deviations for interval data) were determined. Plots of retention relative to time were created for various groups of participants using the Kaplan-Meier method and were formally compared between groups using weighted log-rank tests. To address the hypothesis that a combination of income and other common characteristics of BF outcomes would be associated with retention at 1, 2, 3, 6, 9, 12, 18, and 24 weeks, Cox proportional hazards regression modeling was conducted. Model demographics included income, self-reported race and ethnicity (non-Hispanic black, Hispanic, and a combination of non-Hispanic white and other races [Asian, Native Hawaiian or Pacific Islander, American Indian, and more than one race], age, route of delivery, history of BF, pre-pregnancy body mass index, insurance, return to work, and infant gender.

## Results

### Sociodemographic Characteristics

Due to successful recruitment, in just over 15 months our team increased our target sample to 250 participants representative of greater diversity than Connecticut state population.^39^ Participants (N=250) were self-reported as 118 (47.2%) non-Hispanic white, 43 (17.2%) non-Hispanic Black, 71 (28.4%) Hispanic, and 18 (7.2%) Other. Those who self-identified as Hispanic were 41 (57.7 %) White, 13 (18.3%) Black, 11 (15.5%) more than one race, 3 (4.2%) Native Hawaiian or Pacific Islander, 2 (2.8%) American Indian, and 1 (1.4%) Asian. Other included those who self-identified as the following: 12 (66.7%) non-Hispanic Asian, 5 (27.8%) non-Hispanic more than one race, and 1 (5.6%) non-Hispanic Native Hawaiian or Pacific Islander. Two hundred and forty-four participants reported their yearly income, 89 (36.5%) ≤$50,000 and 155 (63.5%) >$50,000. Other participant characteristics known to affect BF outcomes: age, route of delivery, history of BF, return to work, pre-pregnancy body mass index, education, and infant gender and weight, are reported in Table 1.

**Table 1.** Demographic Characteristics (n=250)

|  | *n* | % |
| --- | --- | --- |
| Age, mean (SD) | 30.24 (4.91) |  |
| Race/Ethnicity |  |  |
| Non-Hispanic White | 118 | 47.2 |
| Non-Hispanic Black | 43 | 17.2 |
| Hispanic | 71 | 28.4 |
| Other | 18 | 7.2 |
| Income |  |  |
| ≤ \$50,000 | 89 | 35.6 |
| > \$50,000 | 155 | 62.0 |
| Missing | 6 | 2.4 |
| History of breastfeeding |  |  |
| Yes | 130 | 52.0 |
| No | 120 | 48.0 |
| Insurance |  |  |
| Medicaid | 91 | 36.4 |
| Private | 139 | 55.6 |
| Self-pay/uninsured_and_other | 14 | 5.6 |
| Missing | 6 | 2.4 |
| Education |  |  |
| Any College or above | 207 | 82.8 |
| High School or below | 43 | 17.2 |
| Infant Gender |  |  |
| Male | 132 | 52.8 |
| Female | 111 | 44.4 |
| Missing | 7 | 2.8 |
| Infant Weight (kg) mean (SD) | 117.182 (28.872) |  |
| Pre-pregnancy BMI mean (SD) | 27.53 (6.45) |  |
| Return to work |  |  |
| ≤6 weeks | 9 | 3.6 |
| >6 weeks | 241 | 96.4 |
| Route of delivery |  |  |
| Vaginal | 167 | 66.8 |
| Cesarean section | 76 | 30.4 |
| Missing | 7 | 2.8 |

### Recruitment and Retention

Retention rates in the study, for 244 women who provided income data, were at 1 (93%), 2 (87%), 3 (85%), and 6 (82%) weeks (Table 2). From weeks 9 through 24, retention rates declined from 77% to 72% (Table 2). Retention over time in the high-income group was significantly higher than in the low-income group (p<0.0001). There was a statistically significant difference in patterns of retention over time between the race and ethnicity categories (p<0.0001). Retention was higher and comparable for non-Hispanic White and Other women, but lower and comparable for non-Hispanic Black and Hispanic women.

**Table 2.** Retention of Key Characteristics at Baseline, 1, 2, 3, 6, 9, 12, 18, and 24 Weeks.

| Variable | Baseline | Week 1 | Week 2 | Week 3 | Week 6 | Week 9 | Week 12 | Week 18 | Week 24 | P value |
| --- | --- | --- | --- | --- | --- | --- | --- | --- | --- | --- |
|  | *n* | *n* | *n* | *n* | *n* | *n* | *n* | *n* | *n* |  |
| Total Sample | 244 | 225 | 211 | 205 | 200 | 187 | 184 | 179 | 175 |  |
| Income |  |  |  |  |  |  |  |  |  | 0.0001 |
| ≤ \$50,000 | 89 | 77 | 73 | 70 | 66 | 56 | 56 | 52 | 50 | |
| > \$50,000 | 155 | 148 | 138 | 135 | 134 | 131 | 128 | 127 | 125 | |
| Race and Ethnicity |  |  |  |  |  |  |  |  |  | 0.0001 |
| Non-Hispanic Black | 40 | 36 | 33 | 30 | 28 | 24 | 23 | 23 | 21 |  |
| Non-Hispanic White | 117 | 111 | 107 | 106 | 103 | 102 | 101 | 98 | 96 |  |
| Hispanic | 70 | 62 | 55 | 53 | 53 | 46 | 45 | 43 | 43 |  |
| Other | 17 | 16 | 16 | 16 | 16 | 15 | 15 | 15 | 15 |  |
| Age mean (SD) | 30.29 (4.94) | 30.4 (4.97) | 30.5 (4.93) | 30.58 (4.9) | 30.66 (4.92) | 30.81 (4.83) | 30.84 (4.75) | 30.92 (4.76) | 30.95 (4.74) | 0.0001 |

To identify individual and combined effects of income and race and ethnicity on retention, Cox Proportional Hazard Regression^40^ analyses were conducted. Due to similarity in retention patterns, the race and ethnicity groups were recoded to a combination of non-Hispanic White and Other women, and a second combination of non-Hispanic Black and Hispanic women. These analyses revealed a hazard ratio (HR) of 2.5 for women of low income compared to those of high income (p=0.0001). This HR implies that low-income status is associated with a 150% increase in the hazard of withdrawal from the study at any point in time. When non-Hispanic Black and Hispanic women were compared to the combined non-Hispanic White and Other group, the HRs were 3.3 and 2.6 respectively (p=0.0001).

In a multivariable Cox model, that included both income and race and ethnicity factors, race and ethnicity continued to be statistically significant (p=0.04), but income was no longer significant (p=0.06). The adjusted HR for low-income women decreased to 1.7. The adjusted HRs for non-Hispanic Black and Hispanic women decreased to 2.4 and 2.0, respectively.

Proportional hazards regression was also used to investigate potential associations between other covariates and retention. For age, the HR was 1.1, when comparing younger women to older women, suggesting that risk of withdrawal increases by 10% with each one year decrease in age (p=0.009). For education, women with attainment of high school or below had a HR of 2.0 compared to those with a college degree or more (p=0.04). Lack of prior BF experience was associated with a HR of 2.6 relative to women with prior BF experience (p=0.051). Other maternal characteristics known to impact BF studies were not significant.

To investigate potential confounding effects, the significant covariates were successively entered into several models that included the retention, income, and race and ethnicity variables. In the model with the income and race and ethnicity factors, none of the covariates remained statistically significant. However, adjustment for age altered the magnitudes of HRs for the income and race and ethnicity effects in a way that suggested it could be a confounding factor. Specifically, the HR for women in the low income versus high income group decreased to 1.6 and HRs for non-Hispanic Black and Hispanic women decreased to 2.1 and 1.9, respectively.

In the final three-factor model of income, self-reported race and ethnicity, and age, the simultaneous statistical test on all three variables was significant (p=.0001). However, none of the individual variables achieved statistical significance. Comparison of nested models with and without age established that, with adjustment for age, the combination of the income and race and ethnicity factors was statistically significant (p = 0.006). This suggests that it is the overlap of information between income and race and ethnicity that results in the model being significantly predictive of withdrawal from the study. More technically, it implies that a collinearity between the three indicator variables for the income and race and ethnicity factors predicts retention as opposed to withdrawal (Table 3).

**Table 3.**
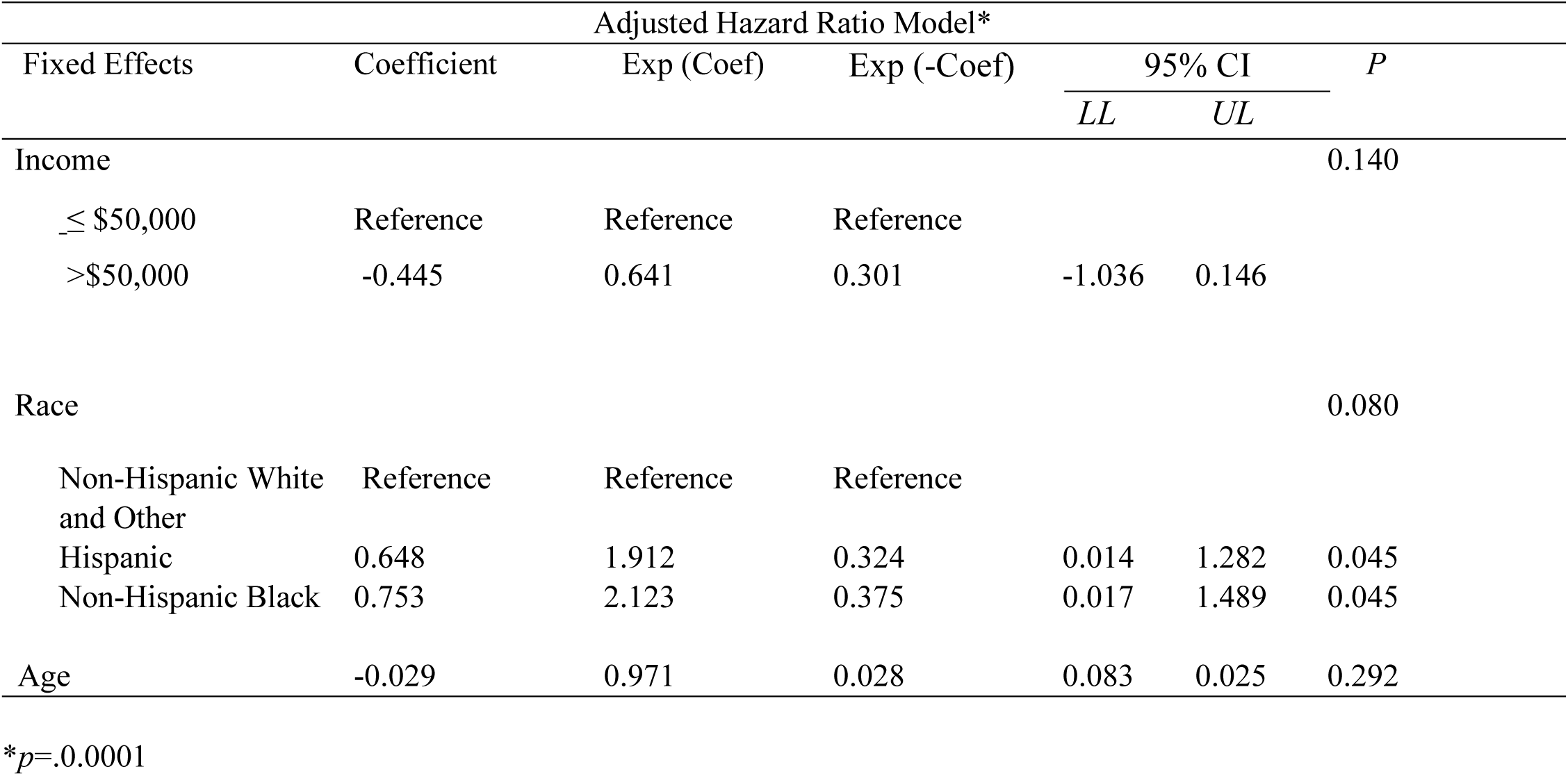
Multiple Cox regression model of characteristics of retention to 24 weeks.

## Discussion

Retention in prospective longitudinal studies of a population below the poverty threshold is challenging in BF and other health-related studies.^23,41^ The present study over-enrolled women from income and racial and ethnic populations who are historically under-represented in clinical studies^28,42^ and are less likely to initiate and sustain BF.^3^ Taken independently, women of low income and diverse race and ethnicity had significantly lower retention in the study. However, when income, race and ethnicity, and age are examined together, their intersection significantly decreases the risk of not being retained in the study. The intersection of these factors represents the structural inequities for women who desire to BF yet are unable to be retained in a BF study.

To facilitate equitable participation, our study utilized an accessible text-based BF intervention to support women to meet their BF goals. Similarly, studies with women below the poverty threshold and Hispanic and non-Hispanic Black women found that the use of a text message–based intervention was acceptable to participants, provided equitable access, and resulted in retention in BF studies.^30,43,44^ Reflective of structural inequities, women in our study who had incomes below the poverty threshold and who self-identified as non-Hispanic Black and Hispanic had a 2.1 and 1.9 times higher risk of not being retained in the study than non-Hispanic White and Other women. Despite inclusive text-based support, women were unable to continue in their chosen BF study.

Historically, women who participate in and complete breastfeeding studies report higher breastfeeding rates on average compared to national rates.^20,44,45^ Furthermore, women below the poverty threshold and who self-identify as non-Hispanic Black and Hispanic are sub-optimally represented in clinical research.^42^ Our study retention reflects these structural inequities, as non-Hispanic Black and Hispanic women had a higher risk of not being retained in the study, with their risk of dropping out not solely accounted for by income. Structural inequities contribute to disparities in BF rates, particularly for non-Hispanic Black and Hispanic women in the US compared with other racial and ethnic groups.^20,46,47^ Cultural values, insufficient BF knowledge and support, and ongoing barriers related to social determinants of health contribute to low initiation of BF for non-Hispanic Black women and early formula introduction among Hispanic women, thus endangering long term BF and impeding retention in a breastfeeding study. ^14,17–20,46^

Factors associated with positive retention in our study were increasing maternal age and previous BF history. In the 2018 CDC National Immunization Survey, age was a significant predictor of breastfeeding behavior, as women ≤ 29 years are less likely to ever breastfeed.^4^ In addition, age and prior BF experience are covariates associated with participation in breastfeeding studies.^4,48^ The intersection of age and previous BF history reflects complex underlying structural inequities which create barriers to Hispanic and non-Hispanic Black women for participation in BF studies and in meeting BF goals.^12,18,49^ Therefore, while text-based BF support is accessible and acceptable, it requires adaptation to address the unique challenges, barriers, and structural inequities encountered by Hispanic and non-Hispanic Black women in meeting their BF goals.^23,41,50,51^

Key to successful retention of participants in our longitudinal study were strategies proven effective in a diverse population.^24,26,45,52^ Skillful implementation of these strategies was driven by our study’s diverse and culturally sensitive research team. Recruitment and face-to-face interviews were conducted by team members hired from the participants’ communities. In-person visits at 6 and 24 weeks were encouraged by flexible scheduling at a time mutually convenient to participant and research team, at the client’s preferred location (hospital or clinic), with easy accessibility to public transportation. Daily assessment of survey completion and follow-up outreach (survey reminder links, and text, telephone, and email messages) were conducted by the project manager to facilitate data collection and participant retention. After observing a pattern of withdrawal after the early weeks of the study, a text message to encourage participant continuation regardless of current feeding method was suggested and implemented. Progressive compensation was also provided to participants, with a bonus for completion.

## Limitations

While this study used an accessible BF intervention, the exclusion criteria of non-English speakers and mothers with infants of ≤ 37 weeks’ gestation may have omitted many women of racial and ethnic diversity who were otherwise eligible. Another limitation was conducting the study in a state with a comparatively high cost of living, reflected in the significant difference in Hispanic and non-Hispanic retention across the study, contributing to BF inequity for women with ≤ $50,000. Beyond this paper’s scope is the effect on retention due to differences in BF pain perception across racial and ethnic groups, the focus of the larger study. To validate the efficacy of a text-based intervention to support BF equity, a larger study, specifically one that includes Spanish-speaking women and mothers of infants of 37 weeks’ gestation, is needed to address underlying structural inequities and provide equitable support for Hispanic and non-Hispanic Black women to reach their BF goals.

## Conclusions

Women of all people groups want to breastfeed. Findings from this study suggest that text-based support is accessible and acceptable for women from diverse income and racial and ethnic backgrounds. However, while women from all income and racial and ethnic groups were successfully enrolled in the current study, Hispanic and non-Hispanic Black women had lower rates of retention, which suggests that there are complex underlying structural inequities, not solely explained by income level. The use of predictor variables, such as age and previous breastfeeding history may inform new strategies for retaining women from lower income and underrepresented racial and ethnic groups in future research of interventions to promote and support breastfeeding. Research should also include collaboration with community partners to aid in identifying and addressing structural inequities to increase retention in future studies and sustain breastfeeding as a positive health outcome.

## Data Availability

All data produced in the present study are available upon reasonable request to the authors

## Acknowledgements

Research reported in this publication was supported by the National Institute of Nursing Research of the National Institutes of Health under Award Number R56NR020041 to Dr. Ruth Lucas. The content is solely the responsibility of the authors and does not necessarily represent the official views of the National Institutes of Health. The authors thank the participants of the PROMPT study for their time and commitment to participation.

## Authorship Contribution Statement

Author 1: Project administration, Writing - review & editing. Author 2: Writing - review & editing. Author 3: Formal analysis, Visualization. Author 4: Writing - original draft. Author 5: Formal analysis. Author 6: Writing - review & editing, Visualization. Author 7: Investigation. Author 8: Project administration. Author 9: Project administration. Author 10: Project administration. Author 11: Writing - review & editing. Author 12: Formal analysis, Writing – review & editing. Author 13: Conceptualization, Methodology, Supervision, Funding acquisition, Writing - review & editing.

## Ethical Conduct of Research

The PROMPT study was performed in line with the principles of the Declaration of Helsinki. Approval was granted by the Ethics Committee of University of Connecticut, Hartford Hospital, and University of Connecticut Health Center (6-30-2021/E-HHC-2021-0143). Informed consent was obtained from all participants in the PROMPT study.

## Clinical Trial Registration

NCT05262920. Registered 2 March 2022. The first participant was enrolled on 16 March 2022. https://clinicaltrials.gov/ct2/show/NCT05262920

## Author Note

**Helen Wilde LaPlant, MS, IBCLC**, is a Project Manager, School of Nursing, University of Connecticut, Storrs, Connecticut, and Program Manager, IBCLC, Hispanic Health Council, Hartford, Connecticut.

**Confidence Francis-Edoziuno, BSN**, is a PhD student and Research Assistant, School of Nursing, University of Connecticut, Storrs, Connecticut.

**Zhe Guan, MA,** is a PhD student and Research Assistant, Department of Statistics, University of Connecticut, Storrs, Connecticut.

**Tumilara Aderibigbe, PhD**, is a Post-doctoral Fellow at University of Utah, Salt Lake City, Utah. At the time this research was completed, she was a Graduate Assistant, School of Nursing, University of Connecticut, Storrs, Connecticut.

**Xiaolin Chang, PhD**, was a Graduate Assistant, Department of Statistics, University of Connecticut, Storrs, Connecticut at the time of data collection.

**Ashwag Saad Alhabodal, RN, MHA/HE**, is a PhD student and Research Assistant, School of Nursing, University of Connecticut, Storrs, Connecticut.

**Kristen Delaney, MS, APRN, FNP-BC**, was a Research Assistant, School of Nursing, University of Connecticut, Storrs, Connecticut at the time this research was completed.

**Dana Scott, MD, FACOG**, is an Assistant Professor, Obstetrics and Gynecology Unit, University of Connecticut Health Center, Farmington, Connecticut.

**Mary Marshall-Crim, MSN, RN, IBCLC**, is Lactation Manager, Women’s Health Services, Lactation Program, Hartford Hospital, Hartford, Connecticut.

**Idelisa Freytes, BSHCA**, is Practice Manager, Women’s Ambulatory Health Services, Hartford Hospital, Hartford, Connecticut.

**Wendy A. Henderson, PhD, CRNP, FAASLD, FAAN**, is a Gail and Ralph Reynolds Presidential Distinguished Professor, Department of Biobehavioral Health Sciences, University of Pennsylvania School of Nursing, and at the time of this research was Director of the PhD Program, School of Nursing, University of Connecticut, Storrs, Connecticut.

**Stephen Walsh, ScD**, is Associate Professor, School of Nursing, University of Connecticut, Storrs, Connecticut.

**Ruth Lucas, Ph.D., RNC, CLS**, **FAAN**, is an Associate Professor, School of Nursing, University of Connecticut, Storrs, Connecticut.

